# Clinical characteristics and prognostic factors of mortality in pediatric patients with biliary atresia awaiting liver transplantation

**DOI:** 10.1101/2025.03.10.25323639

**Authors:** Nguyen Hong Van Khanh, Nguyen Tran Nam Tien, Bui Thanh Liem, Duong Thi Thanh, Le Lam Anh Thy, Truong Thi Yen Nhi, Tran Thanh Tri, Nguyen Phuoc Long, Duc Ninh Nguyen, Bui Quang Vinh

## Abstract

**Background:** Despite advancements in Kasai portoenterostomy (KP) for biliary atresia (BA), 70-80% of patients require liver transplants with long waiting periods time. This study aims to investigate factors associated with mortality in BA patients awaiting liver transplantation.

**Methods:** This cross-sectional study included BA patients indicated for liver transplantation from May 1, 2023 to August 30, 2024. Factors associated with the survival outcome were explored using univariable and multivariable logistic regression. The effect of the KP on overall survival was assessed through Kaplan-Meier survival analysis and multivariable Cox regression.

**Results:** This study included 97 patients with a mortality rate of 16.5%. Multivariable logistic regression showed that KP (adjusted OR (aOR): 0.119; 95% CI: 0.021-0.678; P-value: 0.016), low pediatric end-stage liver disease (PELD) scores (aOR: 1.201; 95% CI: 1.074-1.343; P-value: 0.001), and high weight-for-age (WA) percentiles (aOR: 0.940; 95% CI: 0.884-0.999; P-value: 0.047) were associated with lower mortality. Sub-group analyses in patients with KP also showed low PELD scores (aOR: 1.155; 95% CI: 1.020-1.309; P-value: 0.023) related to lower mortality, whereas high GGT levels showed a potential association with survival (OR: 0.990; 95%CI: 0.981-1.000; P-value: 0.039). Further survival analysis demonstrated the prognostic value of KP (adjusted hazard ratio: 0.261; 95% CI: 0.088-0.770; P-value: 0.015).

**Conclusion:** KP status and PELD scores are significantly associated with mortality risk in BA patients waiting for liver transplants, suggesting their potential role as early indicators for liver transplant prioritization. These findings are significant for informing follow-up care and early intervention strategies, though additional validation is necessary.

## INTRODUCTION

Biliary atresia (BA) is a devastating pediatric condition characterized by inflammatory and fibrotic processes that lead to obstruction of the extrahepatic bile ducts and abnormalities in the intrahepatic bile ducts [1]. BA is rare, with considerably varied incidences across regions globally. In Southeast Asia, it ranges from 1 in 5,000 to 1 in 9,000 live births, while in Europe and North America, it fluctuates from 1 in 15,000 to 1 in 19,000 live births [2]. Upon BA diagnosis, Kasai portoenterostomy (KP) is conducted to replace the obliterated extrahepatic bile duct with the intestinal conduit, thereby restoring bile flow [3]. However, BA is the leading cause of pediatric liver transplantation. Despite the timely intervention and successful outcomes of KP, the necessity for liver transplantation by the age of 20 persists at a rate of 70 to 80%, representing 50% of pediatric liver transplant cases [4]. One-third of children with BA require early liver transplantation within the first 12 to 24 months [5]. However, patients with biliary atresia frequently face prolonged waiting time for liver transplantation [6].

At the two largest pediatric hospitals in southern Vietnam, approximately 46 BA cases are reported annually [7, 8]. The number of patients undergoing KP was 158 over 6.5 years and 65 over 3 years in these two hospitals. At the Children’s Hospital 2, over 90% of liver transplants performed to date have been indicated for underlying BA. Over the past 18 years, our center has conducted liver transplants in only 26 cases, falling short of actual demand. Consequently, a significant number of patients remain on the transplant waiting list. These patients face complications of cirrhosis, such as cholangitis, ascites, and portal hypertension, causing gastrointestinal bleeding, liver dysfunction, malnutrition, hepatopulmonary syndrome, and pulmonary artery hypertension, eventually resulting in an increased risk of mortality [3, 9]. Therefore, identifying risk factors associated with survival outcomes in these patients holds potential for liver transplant prioritization.

This study utilized univariable and multivariable logistic regression with the aim of exploring the risk factors associated with mortality in BA patients waiting for liver transplantation, thereby allowing for timely management. Furthermore, survival analysis was performed to investigate the effect of KP on overall survival. We found that KP status and PELD scores are significantly associated with mortality risk in BA patients awaiting liver transplants, suggesting that those without KP and high PELD scores should be prioritized for liver transplants.

## METHODS

### Study design and study population

The observational study had a protocol approved by the Institutional Review Board of the Children’s Hospital 2, Ho Chi Minh, Vietnam (No. 957/QĐ-NĐ2). We included biliary atresia patients with liver transplant indications, admitted from 01/05/2023 to 31/08/2024. Inclusion criteria were: (1) previously diagnosed with BA, (2) liver transplant indication, (3) currently being followed up at the outpatient setting. Liver transplant indication was defined based on (1) pediatric end-stage liver disease (PELD) score ≥ 10 for patients under 12 years old [10] or model for end-stage liver disease (MELD) score ≥ 15 for patients ≥ 12 years old [11]; (2) portal hypertension with recurrent bleeding complication ≥ 1 time in the last 12 months; or (3) hepatopulmonary syndrome. Patients were excluded if they had long-term contraindications to liver transplantation, defined based on (1) rapidly progressed hepatocellular carcinoma with blood vessel invasion; (2) extrahepatic cancer (except hepatoblastoma with lung metastasis); (3) severe and non-response pulmonary artery hypertension; or (4) other diseases such as mitochondrial diseases with multiple organ damage, Niemann Pick type C disease, and hemophagocytic lymphohistiocytosis.

### Outcome and covariates

The primary outcome was mortality, which was recorded during hospitalization or by phone call after discharge. Covariates considered in the analysis included demographics, history characteristics (obstetrics and biliary atresia history), clinical examination, laboratory tests, cirrhosis severity, biliary atresia complications, and nutrition. We recorded age, sex, birth weight, gestational age, age at biliary atresia onset, history of Kasai portoenterostomy including Kasai portoenterostomy delay time and age at Kasai portoenterostomy, liver transplant history, jaundice, hepatomegaly (categorized as normal liver size, 2 cm, 3 cm, 4 cm, and >4 cm), splenomegaly (categorized as normal spleen size, grade 1, grade 2, grade 3, and grade 4) ascites (categorized as no, mild, and moderate/severe), edema, clubbed fingers, drainage result (categorized as failure and success), white blood cell (WBC), hemoglobin (HGB), platelet count (PLT), aspartate aminotransferase (AST), alanine aminotransferase (ALT), alkaline phosphatase (PAL), gamma-glutamyl transferase (GGT), creatinine, total bilirubin, direct bilirubin, albumin, international normalized ratio (INR), Child-Pugh class (categorized as class A, B, and C), PELD score, portal hypertension, hepatopulmonary syndrome, pulmonary artery hypertension, bleeding, cholangitis, height-for-age (HA) percentiles, WA percentiles, body mass index (BMI) percentiles, acute malnutrition (categorized as no, yes, and severe condition), chronic malnutrition, and vitamin D levels.

### Statistical analysis and modeling

Descriptive statistics were performed to describe the baseline characteristics of the patients corresponding to the survival and non-survival groups [12]. Visual inspections and the Kolmogorov-Smirnov test were used to examine the normality of continuous variables. Mean and standard deviation (SD) were used as summary statistics for variables with normal distribution. The remaining continuous variables were expressed as median and interquartile range (IQR). The number and percentages of patients in each group were reported for categorical variables. Regarding statistical tests, Welch’s two-sample t-test was performed for normally distributed continuous variables, Fisher’s exact test for nominal categorical variables, and Wilcoxon rank sum for non-normally distributed continuous variables or ordinal categorical variables.

Before the logistic regression model development, 4 patients over 12 years old were excluded because the PELD score is not applicable to these patients [13, 14]. Variables with over 20% missing rate were also excluded [15, 16]. Besides, age at Kasai portoenterostomy and Kasai portoenterostomy delay time were also excluded due to only available in the patients who underwent KP. Median imputation and multiple imputation (using *Hmisc* R package version 5.2.0 [17]) were used for continuous variables (birth weight and gestational age) and binary variables (bleeding, cholangitis, and drainage result), respectively. Next, the pair-wise correlation of continuous variables was examined. Among two highly correlated variables (correlation of coefficient, r > 0.7), variables with known clinical values were retained. In order to estimate the odds ratio and 95% confidence intervals (CI), the univariable logistic regression was utilized for all variables. The multivariable logistic regression was first built based on the variables with P-value <0.05 in the univariable logistic regression [18, 19]. Next, backward elimination was carried out to remove variables and build the best model. Nested models were compared via the log-likelihood ratio test. The coefficients estimated in the best model were then employed to derive the adjusted OR and 95%CI. The model building, OR estimates, and 95%CI of OR were conducted in the *finalfit* R package version 1.0.8 [20]. The 95%CI of OR was calculated using the estimates and standard errors of OR as the Wald confidence interval.

A Kaplan-Meier survival analysis and log-rank test were also performed to examine the overall survival differences between patients who underwent the Kasai procedure and those who did not [21]. Time intervals between birth and mortality dates were used to define overall survival. Censoring was labeled to patients who were alive at the study end date. Besides, multivariable Cox regression analysis, adjusting for the variables retained in the best logistic regression model, was performed to estimate the hazard ratio and 95% CI between patients who underwent the Kasai procedure and those who did not. The *survival* R package version 3.7.0 and *survminer* R package version 0.4.9 were used for survival analyses and Kaplan-Meier curves visualization. P-value <0.05 was considered statistically significant. R 4.3.2 was used in all analyses unless otherwise stated [22].

## RESULTS

### Study participants and baseline characteristics

Between May 1, 2023, and August 31, 2024, 99 BA patients were enrolled in this study (**Figure 1**). Survival outcome was missing in two patients due to lost contact during the follow-up period. The demographics and characteristics of the 97 patients, which include 16 non-survivors and 81 survivors, are presented in **Table 1**.

**Figure 1.**
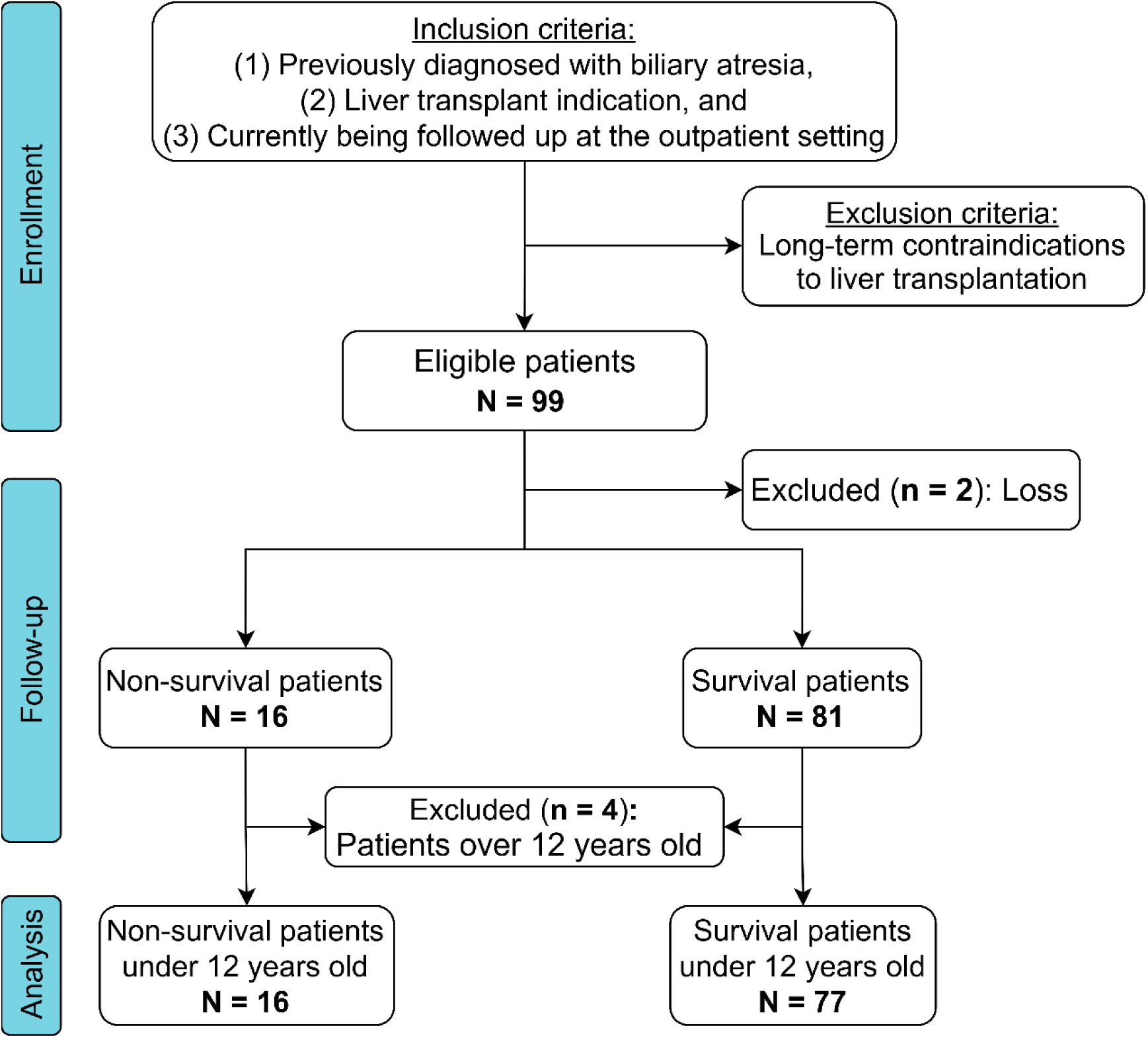
The flowchart of included patients.

**Table 1.** Clinical characteristics between non-survival and survival biliary atresia patients.

| Characteristics | Non-survival<br>(N = 16<sup>1</sup>) | Survival (N = 81<sup>1</sup>) | P value<sup>2</sup> |
| --- | --- | --- | --- |
| Age, days<sup>2</sup> | 256 (199, 577) | 443 (234, 1,233) | 0.052 |
| Sex, male<sup>3</sup> | 9 (56%) | 36 (44%) | 0.4 |
| Birth weight, g<sup>2</sup> | 2,950 (2,600, 3,150) | 3,000 (2,775, 3,300) | 0.3 |
| Missing data | 0 | 1 |  |
| Gestational age, weeks<sup>2</sup> | 39.00 (38.00, 40.00) | 39.00 (38.00, 40.00) | 0.8 |
| Missing data | 0 | 1 |  |
| Age at onset biliary atresia, days<sup>2</sup> | 62 (44, 86) | 50 (30, 75) | 0.2 |
| Kasai portoenterostomy<sup>3</sup> | 9 (56%) | 74 (91%) | 0.002 |
| Kasai portoenterostomy delay, days<sup>2</sup> | 11 (4, 20) | 15 (7, 30) | 0.2 |
| Kasai portoenterostomy age, days<sup>2</sup> | 70 (58, 78) | 74 (59, 90) | 0.4 |
| Liver transplant<sup>3</sup> | 1 (6.3%) | 9 (11%) | >0.9 |
| Jaundice<sup>3</sup> | 16 (100%) | 75 (93%) | 0.6 |
| Hepatomegaly<sup>2</sup> |  |  | 0.6 |
| Normal liver size | 3 (19%) | 12 (15%) |  |
| 2 cm | 4 (25%) | 12 (15%) |  |
| 3 cm | 3 (19%) | 11 (14%) |  |
| 4 cm | 1 (6.3%) | 12 (15%) |  |
| >4 cm | 5 (31%) | 34 (42%) |  |
| Splenomegaly<sup>2</sup> |  |  | 0.079 |
| Normal spleen size | 2 (13%) | 2 (2.5%) |  |
| Grade 1 | 0 (0%) | 15 (19%) |  |
| Grade 2 | 8 (50%) | 32 (40%) |  |
| Grade 3 | 6 (38%) | 25 (31%) |  |
| Grade 4 | 0 (0%) | 7 (8.6%) |  |
| Ascites<sup>2</sup> |  |  | 0.003 |
| No | 6 (38%) | 63 (78%) |  |
| Mild | 3 (19%) | 8 (9.9%) |  |
| Moderate/Severe | 7 (44%) | 10 (12%) |  |
| Edema<sup>3</sup> | 3 (19%) | 5 (6.2%) | 0.12 |
| Clubbed fingers<sup>3</sup> | 9 (56%) | 47 (58%) | >0.9 |
| Drainage result<sup>3</sup> |  |  | 0.019 |
| Failure | 16 (100%) | 58 (73%) |  |
| Success | 0 (0%) | 22 (28%) |  |
| Missing data | 0 | 1 |  |
| WBC, x10<sup>3</sup>/μL<sup>2</sup> | 12.2 (7.5, 14.5) | 9.4 (5.5, 12.4) | 0.11 |
| HGB, g/dL<sup>4</sup> | 9.65 (1.82) | 10.37 (1.74) | 0.2 |
| PLT, x10<sup>3</sup>/μL<sup>2</sup> | 138 (84, 189) | 145 (94, 239) | 0.5 |
| AST, U/L<sup>2</sup> | 250 (154, 304) | 229 (154, 331) | >0.9 |
| ALT, U/L<sup>2</sup> | 133 (85, 170) | 122 (78, 207) | 0.8 |
| PAL, U/L<sup>2</sup> | 421 (306, 569) | 555 (401, 702) | 0.061 |
| GGT, U/L<sup>2</sup> | 88 (45, 194) | 273 (116, 493) | 0.003 |
| Creatinine, μmol/L<sup>4</sup> | 24 (9) | 27 (9) | 0.2 |
| Total bilirubin, μmol/L<sup>2</sup> | 384 (285, 539) | 190 (93, 272) | <0.001 |
| Direct bilirubin, μmol/L<sup>2</sup> | 222 (170, 285) | 128 (63, 190) | <0.001 |
| Albumin, g/L<sup>2</sup> | 23 (20, 26) | 31 (26, 36) | <0.001 |
| INR<sup>2</sup> | 1.88 (1.35, 2.31) | 1.21 (1.10, 1.36) | <0.001 |
| Child-Pugh class<sup>2</sup> |  |  | <0.001 |
| A | 0 (0%) | 5 (6.2%) |  |
| B | 5 (31%) | 65 (80%) |  |
| C | 11 (69%) | 11 (14%) |  |
| PELD score<sup>2</sup> | 26 (21, 28) | 15 (12, 19) | <0.001 |
| Missing data | 0 | 4* |  |
| Portal hypertension<sup>3</sup> | 14 (88%) | 67 (83%) | >0.9 |
| Hepatopulmonary syndrome<sup>3</sup> | 0 (0%) | 3 (3.7%) | >0.9 |
| Pulmonary artery hypertension<sup>3</sup> | 0 (0%) | 1 (1.2%) | >0.9 |
| Bleeding<sup>3</sup> | 2 (13%) | 21 (26%) | 0.3 |
| Missing data | 0 | 1 |  |
| Cholangitis<sup>3</sup> | 6 (38%) | 54 (68%) | 0.045 |
| Missing data | 0 | 1 |  |
| Height-for-age percentiles<sup>2</sup> | 0 (0, 7) | 9 (0, 40) | 0.024 |
| Weight-for-age percentiles<sup>2</sup> | 2 (0, 8) | 10 (2, 43) | 0.003 |
| BMI percentiles<sup>2</sup> | 23 (3, 69) | 44 (12, 69) | 0.3 |
| Acute malnutrition<sup>2</sup> |  |  | 0.008 |
| No | 7 (44%) | 60 (74%) |  |
| Yes | 4 (25%) | 16 (20%) |  |
| Severe | 5 (31%) | 5 (6.2%) |  |
| Chronic malnutrition<sup>3</sup> | 11 (69%) | 32 (40%) | 0.052 |
| Vitamin D, nmol/L<sup>2</sup> | 34 (24, 68) | 55 (22, 83) | 0.5 |

<sup>1</sup>Mean (SD); Median (interquartile range); n (%)
<sup>2</sup>Wilcoxon rank sum test; <sup>3</sup>Fisher's exact test; <sup>4</sup>Welch Two Sample t-test
\*Missing value came from patients over 12 years old
Abbreviations: WBC, White blood cell; HGB, Hemoglobin; PLT, Platelet; AST, Aspartate aminotransferase; ALT, Alanine aminotransferase; PAL, Alkaline phosphatase; GGT, Gamma-glutamyl transferase; INR, International normalized ratio; PELD, Pediatric end-stage liver disease; BMI, Body mass index

The two groups showed no significant differences in age at inclusion, sex ratio, birth weight, and gestational age. The median age at diagnosis of biliary atresia was 62 (IQR: 44, 86) days for the non-survival group and 50 (IQR: 30, 75) days for the survival group. After a median of 11 and 15 days of the BA diagnosis, KP was performed in 9/16 patients and 74/81 patients in the non-survival and survival groups, respectively. The percentage of patients who underwent KP was significantly higher in the survival group (91% versus 56%, P-value = 0.002). None of the non-survived patients had successful drainage results, whereas 28% of survived patients were successful. Ten patients [1/16 (6.3%) in the non-survival group and 9/81 (11%) in the survival group] underwent liver transplants. The proportion of patients with jaundice, hepatomegaly, splenomegaly, ascites, edema, and clubbed fingers was not statistically different between the two groups. Laboratory tests showed statistical differences in GGT, total bilirubin, direct bilirubin, albumin, and INR. Most patients who did not survive were categorized as Child-Pugh C (11/16, 69%), while most survivors were categorized as Child-Pugh B (65/81, 80%). Median PELD scores in the non-surviving group were significantly higher than those in the surviving patients [26 (IQR: 21, 28) versus 15 (IQR: 12, 19), P-value < 0.001]. Regarding nutritional status, the non-survivor group exhibited lower median height-for-age percentiles (P-value = 0.024) and weight-for-age percentiles (P-value = 0.003), along with a higher proportion of patients with acute malnutrition (P-value = 0.008) compared to the survivor group. Median vitamin D levels for the non-survivor and survivor groups were 34 (IQR: 24, 68) and 55 (IQR: 22, 83) nmol/L, respectively.

### Patients with Kasai procedure, lower PELD scores, and higher weight-for-age percentiles were associated with lower odds of non-survival

Univariable logistic regression identified several factors associated with the mortality risk (**Table 2**). Patients who underwent Kasai portoenterostomy (OR 0.129; 95% CI: 0.037-0.452), those with cholangitis (OR 0.288; 95% CI: 0.094-0.883), higher albumin levels (OR 0.841; 95% CI: 0.755-0.936), higher WA percentiles (OR 0.925; 95% CI: 0.863-0.991), and increased GGT levels (OR 0.997; 95% CI: 0.994-1.000) were associated with decreased odds of non-survival. Patients with severe acute malnutrition compared to those without acute malnutrition (OR 8.143; 95% CI: 1.877-35.317), elevated INR (OR 7.145; 95% CI: 1.933-26.416), those experiencing moderate to severe ascites versus those without ascites (OR 6.883; 95% CI: 1.914-24.756), higher PELD scores (OR 1.210; 95% CI: 1.097-1.335), and increased total bilirubin levels (OR 1.008; 95% CI: 1.004-1.012) were associated with an increased odds of non-survival. Total bilirubin, albumin, and INR were excluded from the multivariable logistic regression because these variables were used clinically to calculate PELD scores. They also exhibited a moderate-to-strong correlation with PELD scores with coefficients of correlation of 0.71, 0.47, and 0.67, respectively (**Supplementary Figure S1**). The final multivariable logistic regression model included Kasai portoenterostomy (adjusted OR (aOR) 0.119; 95% CI: 0.021-0.678; P-value = 0.016), PELD scores (aOR 1.201; 95% CI: 1.074-1.343; P-value = 0.001), and WA percentiles (aOR 0.940; 95% CI: 0.884-0.999; P-value = 0.047).

**Table 2.** Estimated odds ratio from univariate and multivariable logistic regression.

|  | Univariate regression |  | Multivariable regression |  |
| --- | --- | --- | --- | --- |
|  | OR (95% CI) | P-value | aOR (95% CI) | P-value |
| Age, days | 0.999 (0.998-1.000) | 0.132 |  |  |
| Sex, Male | 1.714 (0.580-5.253) | 0.331 |  |  |
| Birth weight, g | 0.999 (0.997-1.000) | 0.188 |  |  |
| Gestational age, weeks | 0.933 (0.647-1.372) | 0.714 |  |  |
| Age at onset biliary atresia, days | 1.011 (0.996-1.027) | 0.130 |  |  |
| Kasai portoenterostomy | 0.129 (0.035-0.451) | **0.001** | 0.119 (0.018-0.640) | **0.016** |
| Liver transplant | 0.504 (0.026-2.993) | 0.530 |  |  |
| Jaundice | 9454402.647 (0.000-NA) | 0.993 |  |  |
| Hepatomegaly |  |  |  |  |
| Normal | Ref. |  |  |  |
| 2 cm | 0.889 (0.153-5.544) | 0.895 |  |  |
| 3 cm | 0.727 (0.108-4.851) | 0.735 |  |  |
| 4 cm | 0.222 (0.010-2.089) | 0.226 |  |  |
| >4 cm | 0.392 (0.078-2.222) | 0.259 |  |  |
| Splenomegaly |  |  |  |  |
| Normal | Ref. |  |  |  |
| Grade 1 | 0.000 (NA-2.444e+41) | 0.992 |  |  |
| Grade 2 | 0.267 (0.028-2.499) | 0.219 |  |  |
| Grade 3 | 0.250 (0.025-2.424) | 0.207 |  |  |
| Grade 4 | 0.000 (NA-7.442e+61) | 0.994 |  |  |
| Ascites |  |  |  |  |
| No | Ref. |  |  |  |
| Mild | 3.687 (0.677-17.265) | 0.103 |  |  |
| Moderate/Severe | 6.883 (1.927-25.866) | **0.003** |  |  |
| Edema | 3.323 (0.622-15.328) | 0.128 |  |  |
| Clubbed fingers | 0.866 (0.292-2.655) | 0.796 |  |  |
| Drainage result, Success | 0.000 (NA-1.465e+36) | 0.991 |  |  |
| WBC, x103/μL | 1.067 (0.973-1.172) | 0.156 | | |
| HGB, g/dL | 0.782 (0.551-1.075) | 0.147 |  |  |
| PLT, x103/μL | 0.999 (0.994-1.003) | 0.816 | | |
| AST, U/L | 1.000 (0.997-1.002) | 0.949 |  |  |
| ALT, U/L | 0.999 (0.994-1.003) | 0.695 |  |  |
| PAL, U/L | 0.998 (0.995-1.000) | 0.096 |  |  |
| GGT, U/L | 0.997 (0.993-0.999) | **0.047** |  |  |
| Creatinine, μmol/L | 0.961 (0.890-1.032) | 0.279 | | |
| Total bilirubin, μmol/L | 1.008 (1.004-1.012) | **0.001** | | |
| Direct bilirubin, μmol/L | 1.009 (1.004-1.016) | **0.002** | | |
| Albumin, g/L | 0.841 (0.746-0.927) | **0.002** |  |  |
| INR | 7.145 (2.224-29.689) | **0.003** |  |  |
| Child-Pugh class |  |  |  |  |
| A | Ref. |  |  |  |
| B | 1242171.488 (0.000-NA) | 0.992 |  |  |
| C | 15651360.748 (0.000-NA) | 0.990 |  |  |
| PELD score | 1.210 (1.109-1.353) | **0.001** | 1.201 (1.086-1.364) | **0.001** |
| Portal hypertension | 1.556 (0.375-10.618) | 0.586 |  |  |
| Hepatopulmonary syndrome | 0.000 (NA-7.199e+71) | 0.991 |  |  |
| Bleeding | 0.436 (0.065-1.754) | 0.300 |  |  |
| Cholangitis | 0.288 (0.089-0.864) | **0.029** |  |  |
| Height-for-age percentiles | 0.976 (0.942-1.001) | 0.114 |  |  |
| Weight-for-age percentiles | 0.925 (0.843-0.974) | **0.027** | 0.940 (0.866-0.986) | **0.047** |
| BMI percentiles | 0.992 (0.974-1.008) | 0.345 |  |  |
| Acute malnutrition |  |  |  |  |
| No | Ref. |  |  |  |
| Yes | 2.171 (0.514-8.226) | 0.262 |  |  |
| Severe | 8.143 (1.864-37.111) | **0.005** |  |  |
| Chronic malnutrition | 3.094 (1.020-10.626) | 0.054 |  |  |
| Vitamin D, nmol/L | 0.994 (0.981-1.004) | 0.313 |  |  |

Abbreviations: OR, odds ratio; aOR, adjusted odds ratio; ref, reference; NA, not applicable; WBC, White blood cell; HGB, Hemoglobin; PLT, Platelet; AST, Aspartate aminotransferase; ALT, Alanine aminotransferase; PAL, Alkaline phosphatase; GGT, Gamma-glutamyl transferase; INR, International normalized ratio; PELD, Pediatric end-stage liver disease; BMI, Body mass index

Focusing on patients who underwent Kasai portoenterostomy, we performed the same univariable and multivariable logistic regression analyses. The OR and aOR are presented in **Table 3**. In the univariable analysis, patients with higher albumin, higher PAL, and higher GGT levels exhibited lower odds of non-survival. Additionally, patients with moderate to severe ascites, elevated WBC, higher total bilirubin, increased INR, and higher PELD scores were associated with greater odds of non-survival. The final multivariable logistic regression model included PELD scores (aOR 1.155; 95% CI: 1.020-1.309; P-value: 0.023) and GGT (aOR: 0.995; 95% CI: 0.988-1.003; P-value: 0.22).

**Table 3.** Estimated odds ratio from univariable and multivariable logistic regression in patients with Kasai portoenterostomy.

|  | Univariable logistic regression |  | Multivariable logistic regression |  |
| --- | --- | --- | --- | --- |
|  | OR (95% CI) | P-value | aOR (95% CI) | P-value |
| Age, days | 0.999 (0.998-1.000) | 0.301 |  |  |
| Sex, Male | 1.067 (0.264-4.314) | 0.928 |  |  |
| Birth weight, g | 0.999 (0.997-1.001) | 0.486 |  |  |
| Gestational age, weeks | 0.991 (0.613-1.603) | 0.972 |  |  |
| Age at onset biliary atresia, days | 0.993 (0.964-1.022) | 0.638 |  |  |
| Liver transplant | 0.847 (0.095-7.595) | 0.882 |  |  |
| Jaundice | 5890820.174 (0.000-Inf) | 0.993 |  |  |
| Hepatomegaly |  |  |  |  |
| Normal | Ref. |  |  |  |
| 2 cm | 2.545 (0.234-27.709) | 0.443 |  |  |
| 3 cm | 1.556 (0.116-20.854) | 0.739 |  |  |
| 4 cm | 0.636 (0.034-11.909) | 0.762 |  |  |
| >4 cm | 0.219 (0.012-3.936) | 0.303 |  |  |
| Splenomegaly |  |  |  |  |
| Normal | Ref. |  |  |  |
| Grade 1 | 0.000 (0.000-Inf) | 0.995 |  |  |
| Grade 2 | 0.345 (0.026-4.557) | 0.419 |  |  |
| Grade 3 | 0.316 (0.021-4.660) | 0.401 |  |  |
| Grade 4 | 0.000 (0.000-Inf) | 0.996 |  |  |
| Ascites |  |  |  |  |
| No | Ref. |  |  |  |
| Mild | 5.238 (0.742-36.980) | 0.097 |  |  |
| Moderate/Severe | 9.167 (1.725-48.722) | **0.009** |  |  |
| Edema | 2.062 (0.205-20.797) | 0.539 |  |  |
| Clubbed fingers | 1.333 (0.308-5.776) | 0.701 |  |  |
| Drainage result, Success | 0.000 (0.000-Inf) | 0.994 |  |  |
| WBC, x103/μL | 1.143 (1.010-1.294) | **0.035** | | |
| HGB, g/dL | 0.671 (0.425-1.059) | 0.087 |  |  |
| PLT, x103/μL | 0.999 (0.993-1.005) | 0.745 | | |
| AST, U/L | 0.997 (0.991-1.003) | 0.296 |  |  |
| ALT, U/L | 0.997 (0.989-1.005) | 0.462 |  |  |
| PAL, U/L | 0.994 (0.989-0.999) | **0.020** |  |  |
| GGT, U/L | 0.990 (0.981-1.000) | **0.039** | 0.995 (0.988-1.003) | 0.220 |
| Creatinine, μmol/L | 0.957 (0.866-1.058) | 0.390 | | |
| Total bilirubin, μmol/L | 1.006 (1.002-1.011) | **0.008** | | |
| Albumin, g/L | 0.792 (0.678-0.926) | **0.003** |  |  |
| INR | 8.200 (1.817-37.011) | **0.006** |  |  |
| Child-Pugh class |  |  |  |  |
| A | Ref. |  |  |  |
| B | 1467062.522 (0.000-Inf) | 0.995 |  |  |
| C | 33090410.211 (0.000-Inf) | 0.994 |  |  |
| PELD score | 1.216 (1.083-1.367) | **0.001** | 1.155 (1.020-1.309) | **0.023** |
| Portal hypertension | 1.825 (0.209-15.890) | 0.586 |  |  |
| Hepatopulmonary syndrome | 0.000 (0.000-Inf) | 0.995 |  |  |
| Bleeding | 0.964 (0.182-5.110) | 0.966 |  |  |
| Cholangitis | 0.800 (0.182-3.513) | 0.768 |  |  |
| Height-for-age percentiles | 0.950 (0.890-1.015) | 0.128 |  |  |
| Weight-for-age percentiles | 0.938 (0.869-1.013) | 0.102 |  |  |
| BMI percentiles | 0.995 (0.974-1.017) | 0.646 |  |  |
| Acute malnutrition |  |  |  |  |
| No | Ref. |  |  |  |
| Yes | 2.446 (0.517-11.578) | 0.259 |  |  |
| Severe | 2.650 (0.246-28.502) | 0.421 |  |  |
| Chronic malnutrition | 2.828 (0.653-12.239) | 0.164 |  |  |
| Vitamin D, nmol/L | 0.998 (0.987-1.010) | 0.759 |  |  |

Abbreviations: OR, odds ratio; aOR, adjusted odds ratio; ref, reference; NA, not applicable; WBC, White blood cell; HGB, Hemoglobin; PLT, Platelet; AST, Aspartate aminotransferase; ALT, Alanine transaminase; PAL, Alkaline phosphatase; GGT, Gamma-glutamyl transferase; INR, International normalized ratio; PELD, Pediatric end-stage liver disease; BMI, Body mass index

### Patients with Kasai procedure were related to higher overall survival

In the survival group, patients with KP had a higher age than those without KP (**Figure 2**). The Kaplan–Meier survival curve indicated a statistically significant improvement in overall survival for patients who underwent KP compared to those who did not (log-rank P-value < 0.0001). Specifically, the survival probability for patients who received KP was 91.54% in year 1, declined to 88.92% in year 2, and dropped to 79.98% in year 15 (**Figure 3**). In patients who did not undergo KP, the survival probability was 66.67% in year 1 and further decreased to 22.67% in year 3. The estimated HR from univariable Cox regression was 0.117 (95% CI: 0.041-0.336). Multivariable Cox regression analysis, adjusted for PELD score and WA percentiles, also demonstrated that patients with KP were associated with a lower risk of fatal outcomes, with an adjusted HR of 0.261 (95% CI: 0.088-0.770; P-value = 0.015).

**Figure 2.**
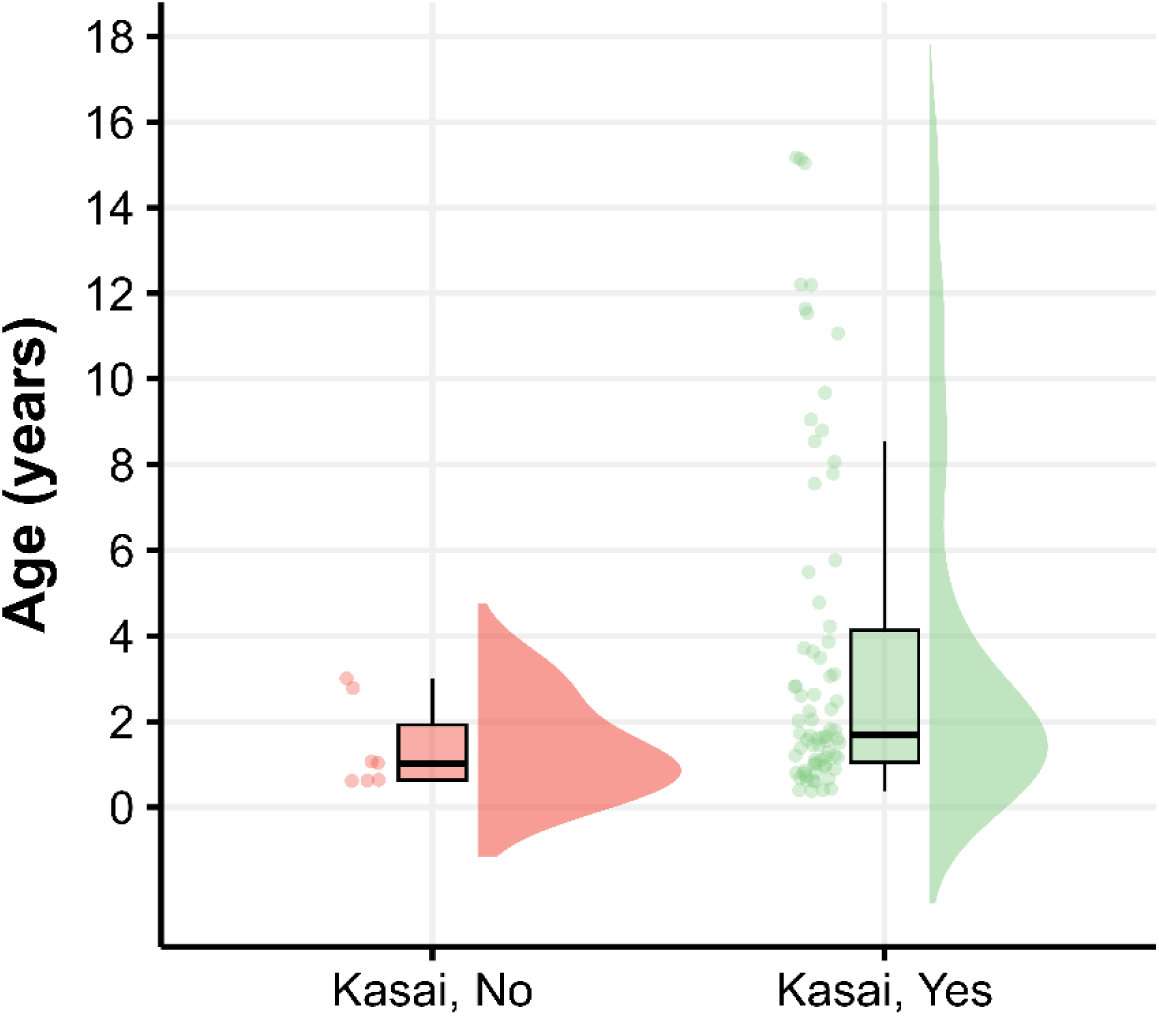
Age distribution at the end of the study in the survival group.

**Figure 3.**
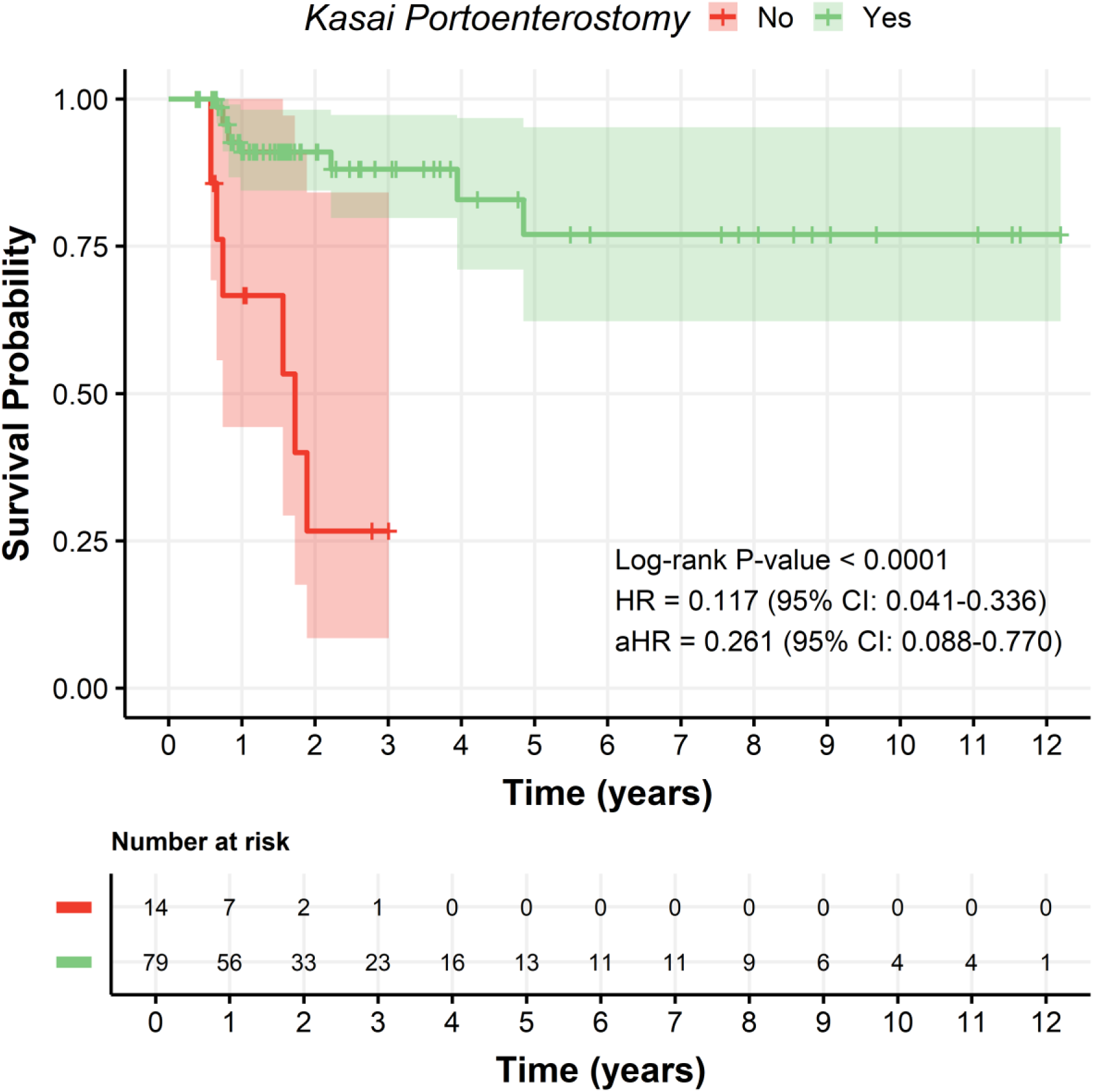
Kaplan–Meier estimates of the overall survival of biliary atresia patients stratified by Kasai portoenterostomy. Shaded areas indicate 95% confidence intervals. HR: Hazard ratio and aHR: adjusted HR (controlled for pediatric end-stage liver disease score and weight-for-age percentiles).

## DISCUSSION

Our study explored risk factors associated with mortality in 97 BA patients awaiting liver transplantation. Our findings indicate that KP, lower PELD scores, and higher WA percentiles were related to decreased mortality. Lower PELD scores remained a significant protective factor among patients who underwent KP, whereas higher GGT levels showed a potential association with survival.

The protective role of KP in delaying liver transplantation and improving survival has been well established. Our study confirmed that patients who underwent KP had significantly better overall survival, consistent with reports from large cohorts in Asia and Europe [23, 24]. In contrast to previous studies, we did not reveal a significant association between the age at KP and mortality outcomes. This discrepancy may be due to the limited sample size or the relatively short follow-up period.

The PELD score has been widely validated as a predictor of pre-transplant mortality in pediatric liver disease [25]. Our study supports its significance, demonstrating a 20.1% increase in mortality risk per unit increase in PELD score. A large study of 4,298 patients with cholestatic liver disease awaiting transplantation demonstrated the accuracy of the PELD score in estimating pre-transplant mortality risk. This study found that 90-day mortality estimates based on PELD scores underestimated actual mortality rates by up to 17% [25]. Therefore, our findings highlight the critical need for early transplant listing, particularly in patients experiencing rapidly progressive liver dysfunction.

Malnutrition has been recognized as a major prognostic factor in BA [26]. Our study identified WA percentiles as an independent predictor of survival, with severe acute malnutrition significantly associated with mortality. These findings align with current literature that emphasize the impact of nutritional status on pre- and post-transplant outcomes [26].

GGT is a controversial factor in the prognosis of diseases, including hepatobiliary diseases. A large study in Korea revealed that higher serum GGT levels were significantly associated with increased mortality risk from cardiovascular disease, cancer, respiratory disease, and liver disease [27]. However, a large study of 1998 BA patients in China found that patients with high GGT levels had a higher two-year survival rate compared to those with low GGT levels (66.3% versus 52.5%; P-value < 0.001). Furthermore, GGT was also identified as an independent predictor of native liver survival (P-value < 0.001, HR = 1.02, 95% CI: 1.01–1.03) in this study. Low GGT levels are seen in other pediatric cholestatic liver diseases and are associated with more severe liver damage. Some BA patients may present with normal or slightly increased GGT, causing delays in diagnosis and Kasai surgery, which can impact surgical success and long-term liver function [28]. In our study, low GGT was found to be associated with mortality.

This study has some limitations. First, the observational and single-center design may limit the generalizability of our findings. Second, our follow-up period was relatively short, preventing long-term survival analysis post-transplant. Future studies with extended follow-up would help refine risk stratification. Third, while we identified key prognostic factors, other clinical parameters, such as specific nutritional interventions or management strategies for portal hypertension, were not evaluated in depth. Further multicenter prospective studies incorporating these factors could provide a more comprehensive understanding of BA outcomes.

## CONCLUSION

Our study highlights the key factors associated with mortality in BA patients awaiting liver transplantation. Kasai portoenterostomy significantly improves survival, while higher PELD scores and lower WA percentiles predict increased mortality risk. These findings emphasize the need for early surgical intervention, optimized nutritional management, and timely liver transplantation to improve outcomes in this high-risk population.

## CRediT authorship contribution statement

**Nguyen Hong Van Khanh**: Conceptualization, Data curation, Methodology, Investigation, Formal analysis, Validation, Writing – Original Draft, Writing – Review & Editing. **Nguyen Tran Nam Tien**: Data curation, Methodology, Investigation, Formal analysis, Validation, Writing – Original Draft, Writing – Review & Editing. **Bui Thanh Liem**: Data curation, Methodology, Investigation, Formal analysis, Supervision, Writing – Review & Editing. **Duong Thi Thanh**: Data curation, Methodology, Validation, Writing – Review & Editing. **Le Lam Anh Thy**: Data curation, Methodology, Validation, Writing – Review & Editing. **Truong Thi Yen Nhi**: Data curation, Methodology, Validation, Writing – Review & Editing. **Tran Thanh Tri**: Data curation, Methodology, Validation, Writing – Review & Editing. **Nguyen Phuoc Long**: Data curation, Methodology, Investigation, Formal analysis, Validation, Supervision, Writing – Review & Editing. **Duc Ninh Nguyen**: Data curation, Methodology, Investigation, Formal analysis, Validation, Supervision, Writing – Review & Editing. **Bui Quang Vinh**: Conceptualization, Data curation, Methodology, Investigation, Validation, Resources, Supervision, Writing – Review & Editing.

## Data availability statement

The data supporting this study’s findings are available upon reasonable request.

