## Supplementary Materials for "Clinical characteristics and prognostic factors of mortality in pediatric patients with biliary atresia awaiting liver transplantation"

### Supplementary Figures

**Figure S1. Correlation between continuous variables.**

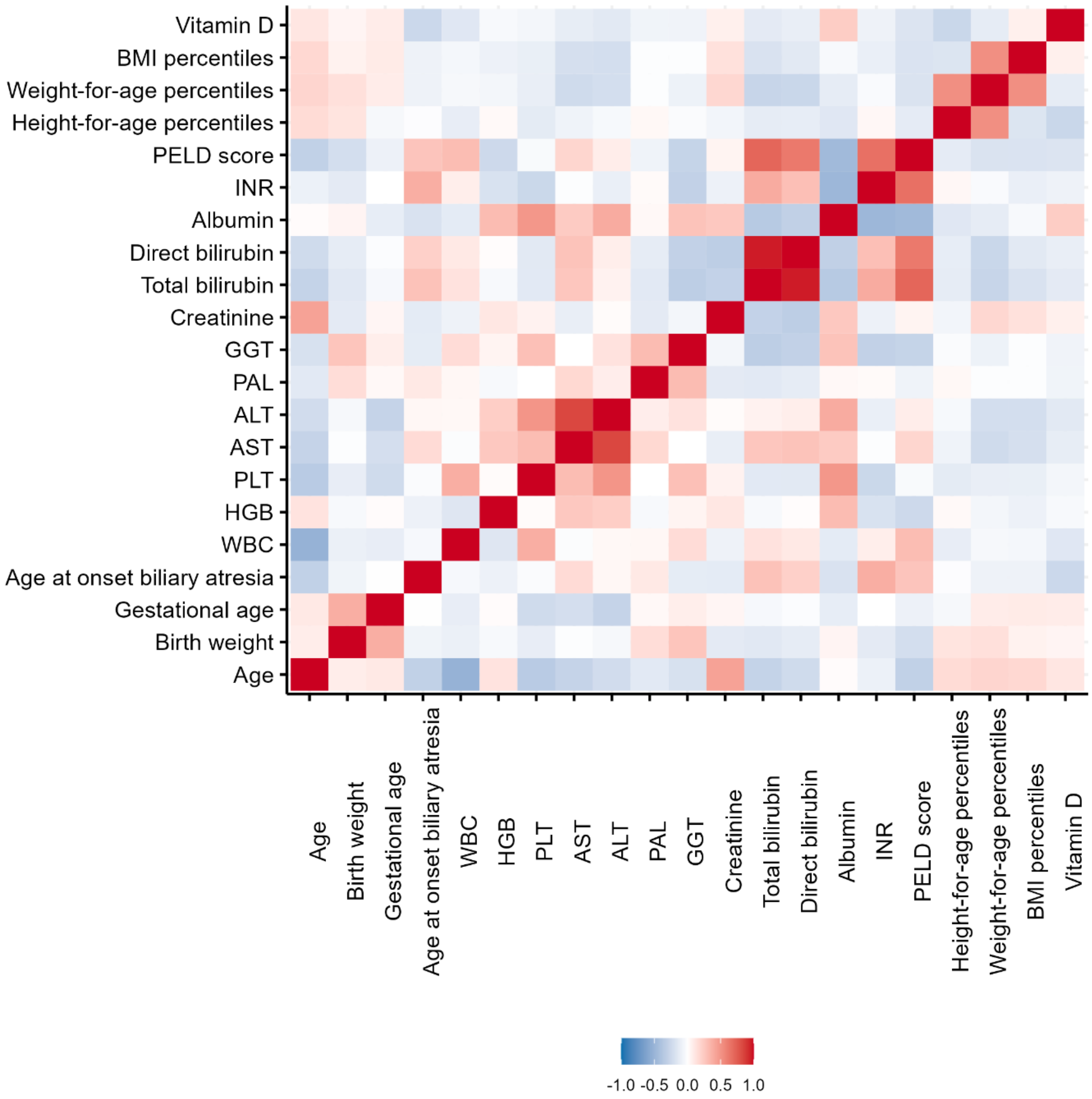
